# Lessons learnt from community referral and follow up of sick young infants with Possible Severe Bacterial Infection in Turkana County, Kenya

**DOI:** 10.1101/2022.08.25.22279210

**Authors:** Samuel Mbugua, Peter Mwaura, Laura Oyiengo, Wilson Liambila, Andrew Emuria, Daniel Gatungu, Jesse Gitaka

**Affiliations:** Directorate of Research and Innovation, Mount Kenya University, 342-0100, Thika, Kenya; Ministry of Health, Kenya; Population Council Kenya, Avenue 5, 3rd Floor, Rose Avenue, PO Box 17643-00500, Nairobi, Kenya; Ministry of Health, Turkana County, Kenya

**Keywords:** Community referral, follow up, young infants

## Abstract

**Introduction:** Management of possible severe bacterial infections in young infants (0-59 days) requires timely identification of danger signs and prompt administration of efficacious antibiotic treatment. The Possible Severe Bacterial Infection guidelines underscore the importance of close follow up in an outpatient basis to ensure treatment adherence and early detection of illness-related complications. The purpose of this study was to strengthen the follow up and referral of sick young infants on day 4 and 8 by introducing community-led interventions that facilitated community health volunteers to identify sick young infants, conduct community reviews, link data with responsive facilities, and refer appropriately.

**Methods:** Six health facilities were included a a longitudinal, descriptive, mixed methods approach weaved around an initial formative context assessment and three-monthly assessments. Quantitative data was extracted from facility registers to identify gaps in follow up and referral feasibility. Qualitative data was through focus group discussions with community health volunteers and key informant interviews with frontline providers.

**Results:** Qualitative data provided insights into key barriers and facilitators of community follow up and referral. Barriers include community socio-cultural practices, competing tasks, dysfunctional community referral pathway, drivers of common infections, and unavailability of essential commodities. Key facilitators entail indication of competency in identification of danger signs in sick young infants, presence of older women, men, and community resource persons that can leveraged on in community engagement and sensitization, and mothers are the primary decision makers in care seeking. There was increased utilization of decision support tools and an increase in the number of sick young infants managed in dispensaries. The COVID-19 pandemic however negatively impacted community follow up and referral of sick young infants.

**Conclusion:** This study seeks to contribute evidence on strengthening PSBI community management by enhancing day 4 and day 8 follow up, review and community referral of sick young infants in Turkana, Kenya. The feasibility, adoption, and fidelity of strengthening community facility linkage through integrated communication strategies was documented, indicative of a successful community-facility linkage in dispensaries and health centers despite the effects of the COVID-19 pandemic.

## Introduction

Severe bacterial infections are responsible for 25% of all neonatal fatalities worldwide [1-4]. In 2018, there were 4 million infant deaths [5]. Africa and South Asia have context-specific inadequacies in service readiness and compromised quality of care in maternal and child health [1,6-8] resulting in various challenges in primary health care provision.

Death from Possible Severe Bacterial Infections (PSBI) is preventable with prompt administration of efficacious antibiotic treatment [9]. The recommended management of sick young infants with PSBI is inpatient treatment (seven to ten days antibiotic course of benzylpenicillin or ampicillin and gentamicin). Many young infants with signs of PSBI do not receive the appropriate care due to inaccessibility, unaffordability, and unacceptability of health services by caregivers and the family [9,10]. The 2015 World Health Organization (WHO) PSBI guidelines were developed by a panel of international experts in child health following clinical trials in Africa and Asia [11,12]. The clinical trials provided evidence of efficacy on the use of simplified treatment regimen for management of PSBI. The guidelines are intended for use in low resource settings where referral of sick young infants is not feasible and do not replace the WHO recommendation on inpatient management as the preferred treatment option for sick infants with clinical severe infection and critical illness [9,11]. Underscored in the guidelines is the importance of close follow up in an outpatient basis to ensure treatment adherence and early detection of illness-related complications. A continuum of care approach envisioned in the implementation of PSBI guidelines entails early identification of danger signs, timely care seeking and treatment, and the back-referral to the community for follow up to ensure treatment completion [13].

Postnatal home visits are essential in prevention and early identification of newborn and young infant health challenges inclusive of syndromic sepsis [14]. The WHO recommends home visits by health workers and community health workers to accelerate early identification of danger signs and promotion of positive care seeking [15,16]. In the context of health systems in low-resource settings, it necessitates the use of community health workers (CHWs) and community health volunteers (CHVs) within the community health system. Early postnatal visits provide a chance for health personnel to provide preventive and promotive care to caregivers. In view of PSBI, the visits by CHVs provide a unique opportunity for the identification of danger signs and appropriate referral of sick young infants [14].

Day 8 follow up visit is essential for assessment of treatment compliance and completion, and review of outcomes of infants on the simplified treatment regimen [17]. Lack of compliance to referral from the community to facilities for sick young infants and follow up on days 4 and 8 as per the WHO 2015 PSBI guidelines [9] has been noted to be a major impediment to quality of care for sick young infants. Poor referral pathways from the community to responsive health facilities worsens care seeking delays.

A system of maintaining records and information is mandatory as the review on day 4 and 8 and referral play an important role in the monitoring and evaluation of performance indicators of the healthcare system. We sought to strengthen the follow up and referral of sick young infants on day 4 and 8 by introducing community-led interventions that facilitated CHVs to identify sick young infants, conduct community reviews, link data with responsive facilities, and refer appropriately. This paper provides evidence and the lessons learnt on the feasibility of integrating community referral and follow up continuum care approach in a low-resource setting whilst adapting PSBI guidelines in Turkana, Kenya.

## Materials and methods

### Context

Our study sites were six health care facilities, and the communities they serve, offering primary care in Turkana central sub-county, Turkana County. The six facilities included one county referral hospital, two health centres and three dispensaries. Turkana central sub-county was part of the Ponya Mtoto project that contributed to reduction in young infant deaths from PSBI. The project was implemented by a consortium of organizations that included Population Council, Mount Kenya University and Kenya Paediatric Research Consortium (KEPRECON). Kenya’s ministry of health has a four-tier system for delivery of health services and follows the Integrated Management of Newborn and Childhood and Illnesses (IMNCI) protocols for management of sick young infants in primary health facilities. Each facility has at least one community unit attached to it comprised of a CHW leading a team of 8-12 CHVs. The frontline health providers hold the responsibility of assessing, classifying, and treating young infants according to the PSBI guidelines. The CHVs’ mandate is assessing a young infant in the community during household visits and prompt referral for further management in the health facility.

### Research design and data collection

We employed a longitudinal, descriptive, mixed methods approach weaved around an initial formative context assessment that assessed the barrier and facilitators to follow up and community referral. This informed the implementation process by informing the development of integrated communication strategies and documentation of outcomes. 3 monthly follow up assessments were conducted to assess the adoption and feasibility of strengthened communication and facility linkage. The target population was frontline health providers and community health volunteers implementing PSBI guidelines in the participating counties. Data collection entailed assessing barriers/bottlenecks and facilitators to day 4 and 8 review, follow up and community referral through qualitative methods; focus group discussions with community health volunteers, key informant interviews with frontline providers and observation field notes by CHEWs. Quantitative data was collected from from secondary data sources (facility registers) to identify gaps in referral feasibility and follow up.

### Data Analysis

Qualitative analysis involved quality control where 10% of the sample was checked to ensure consistency between the transcripts and audios recordings. A thematic framework was developed using an iterative approach and used to classify and organize data into key themes/ constructs. Second iterative reading refined the emergent themes. Coding and recoding of data was performed using Nvivo 12. Emergent categories were identified during analysis. Analysis charts were prepared for each theme and category of participants and used to identify common themes across participants and sites.

Quantitative data descriptive analysis was analysed using STATA version 17 (Statacorp LP) AND entailed standard deviation, means and interquartile range. We estimated the percentage of sick young infants with PSBI identified by CHVs and at service delivery points are based on expected annual births in the two sub counties.

### Ethical considerations

Ethical clearance was sought from MKU-Institutional Scientific Ethics Review Committee MKU/ERC/1596. All the study activities and data handling were in strict compliance to national and international ethical standards. Written informed consent was obtained from each participant before conducting an interview. Personal identifiers were not included during data collection, data entry or analysis.

### Project Implementation

The implementation research study was embedded in the Ponya Mtoto project that sought to provide implementation evidence of contextual scale up of implementation of simplified antibiotic regimens for sick young infants 0-59 days with PSBI in Kenya. The project sought to contribute to the reduction of deaths of young infants in Kenya from possible severe bacterial infection (PSBI) through adaptation of the WHO PSBI guidelines and scaled up implementation within the revised national IMNCI guidelines. The project sought to address Kenya’s high neonatal mortality rate which is at 22 per 1000 live births with sepsis contributing up to 20% [18, 19]. Project activities were implemented by the Ministry of Health through the county, Sub County and health facility management teams and committees.

Implementation of PSBI guidelines leverages on the community health system spearheaded by CHVs who offer services directly to individual households. The CHVs are tasked with follow up where referral is not feasible and community referral of sick young infants to health facilities.

## Results

### 1. Objective 1: Barriers and facilitators of a community centric PSBI management approach

#### Facilitators

##### 1.1. Competency in identification of danger signs in sick young infants by Community Health Volunteers (CHVs)

Focus group discussions with CHVs indicated general competency in identification of danger signs of possible severe bacterial infection in sick young infants. This is evident in the qualitative narratives below;

> *“The first thing we look out for is the child’s overall health. If the child’s health seems okay and he/she is stable, you can safely say that the child is fine. But if the child’s health is declining, this is a clear sign that the child is sick. So you have to take an action of scrutinizing how to help the child. The first step is to seek information from the child’s parents and after discovering the child’s problem, you’re obligated to arrange for the child’s treatment. For example, as CHVs (Community Health Volunteers), we are mandated to write the child a referral so he/she visits a doctor to get the necessary treatment.” [****CHV in Focus Group]***
>
> *“One of the danger signs we look out for is a child with high fever and the child is not breast feeding, vomiting, and diarrhea. We then refer the child and ask the mother to take the child to hospital.”* ***[CHV in Focus Group***]

Some CHVs indicated that they felt undermined in their duties and responsibilities pointing at a perception of inadequacy in their roles from the community;

> *“For instance, the community is well aware that we don’t treat the children so sometimes parents will take their children to hospital without consulting us first. So they undermine us because all we’re good for is simply writing a referral for the child. So we rarely know the number of children that become sick and you only get to know when the child arrives at the hospital displaying symptoms such as fast breathing which we are taught to manage.”* ***[CHV in Focus Group***]

##### 1.2. Community platforms to strengthen young infant care

Some community-based platforms and low lying fruits that were identified and can be leveraged on in strengthening community referral and follow up include community engagement forums (barazas) for engagement and health education, regular meetings between CHVs and health providers to strengthen the community system linkage, county support and motivation of CHVs.

Community engagement forums:

> *“One is to set up a baraza (public meeting) within the village…*..*Also, CHVs conduct health education and in doing so, they educate the community on the significance of immunization.”*
>
> [***CHV in Focus Group***]

Regular meetings with CHVs and facilitation of referral:

> *“Okay in regards to CHVs, I think we should have a meeting, ask their initials, so that we get to know them so that it will improve communication between us and them and our patients. In terms of patients, if we communicate more with our patients and also some kind of support in terms of reimbursement because we talked about transport which means finances-so if some reimbursement is introduced, it will also be an encouragement to the patients and also proper communication.” [****Provider KII]***

County support and motivation of CHVs:

> *“So I would encourage the county to support CHVs like the companies (NGOs) used to and at least allow us once again to go for dialogue days once a month so we are able to educate pregnant mothers to come to hospital which I think will improve the situation.” [****CHV in Focus Group***]

Providers opined that the inclusion of a referral note, education and sensitization, follow up and review on day 4 and 8 was feasible. This are significant strategies that can be leveraged on to ensure treatment completion and the fidelity of day 4 and 8 follow up.

> *“….my opinion is if I give a referral note to the caregiver to go home with it, the CHV from that area should come to review the child the fourth day. They should encourage the mother to bring the child to the hospital.” [****Provider KII***]
>
> *“Education of CHVs and sensitizing the community on these sicknesses of the young infants so that they also know some illnesses can be cured without the intervention of traditional cures.” [****CHV in Focus Group***]

Prioritization of sick young infants in facility care provision was mentioned as a key strategy employed to ensure timely care provision. This meant that caregivers did not have to queue with other patients seeking health services in the facility.

> *“And what we have instructed those outside (caregivers) that for a child below two months, bring them and pass everybody. So they should not delay anywhere.” [****Provider KII***]

##### 1.3. Role of older women, men, and community resource persons

The elders (men and women), also referred to as community resource persons, are instrumental in community sensitization and engagement.

> *“…. the chief might be handed information to pass on to people and the community gets to know about new information regarding the caring for the health of children. The chiefs and elders’ role is therefore limited to disseminating information through barazas. [****CHV in Focus Group***]

Local healers, the village chiefs, and grandmothers were some of the identified community resource persons that can be leveraged on in the care of sick young infants.

> *“For instance local healers work in collaboration with the village chief so if there is a case involving a sick child, they will notify us.” [****CHV in Focus Group***]

##### 1.4. Decision making in care seeking

The primary decision maker when seeking care for sick young infants is the mother. The CHVs also have a significant role in initiating community referral to the health facility on identification of a sick infant,

> *“More often than not, mothers are the decision makers regarding the wellbeing of the child. Men on the other hand are in most instances absent from the home or simply watch from a distance. Church men are the exception but all in all the mothers are the ones who rush the child to hospital.” [****CHV in Focus Group***]

#### Barriers

##### 1.5. Community socio-cultural practices resulting in PSBI

Several socio-cultural practices were identified as having a correlative causal effect to PSBI in young infants. These include naming ceremonies which resulted in nutritional deficiencies associated with delays in initiation of breastfeeding, removal of teeth culturally interpreted as the cause of diarrhea, application of ash as medication resulting in cord sepsis among others. Examples of narratives indicative of these practices are highlighted below.

Naming ceremonies:

> *“In other instances, during childbirth, there are practices that guide the naming of a child. The child is not allowed to breastfeed on the spot. We’ve explained to them that a baby should be allowed to breastfeed within the shortest time possible after birth. Even so, people here wait until the next day so the child is given a name a first before they are breast fed. This practice is harmful to the child.”* .*”[* ***CHV in Focus Group***]

Removal of teeth:

> *“…*..*For the teeth extraction for example, it is done with a nail. A single nail is used on multiple kids…*.. *Sometimes they even use their nails to press what they term as plastic teeth. Then for the more developed teeth, they consider them suitable for extraction with a nail.” [****CHV in Focus Group***]

Application of ash:

> *“….some still believe in the cultural approach of applying ash which is perceived to work quicker than the hospital medicine.”[* ***CHV in Focus Group]***

Some key social issues emergent indicated at a larger societal problem requiring a wider scope of approach in assessing and developing contextual interventions. These included alcoholism with poverty as key predisposing factor.

> *“….parents are unable to buy prescribed medication at the chemist, sometimes due to poverty, sometimes due to the parent’s alcoholism. The parent is less likely to purchase the prescribed drugs upon leaving the hospital.” [****CHV in Focus Group***]

##### 1.6. Competing tasks

Some mothers are sometimes faced with conflicting roles that affected decision making to seek care for their sick infants. The key contributing factors towards this was employment and poverty.

> *“Some women lack the financial ability to bring their child to the facility. Sometimes the mother has other children at home which makes it difficult for her to spend a night with the sick child at the hospital. Moreover, the mother will also get discouraged by the fact that inpatient services here at the facility cost money which they don’t have. This is why some mothers simply want their child to receive treatment and take the child with them back home on the same day.” [****CHV in Focus Group***]

##### 1.7. Dysfunctional community referral pathway

Functional referral pathways existed in most facilities. There are however significant deficits, across all facilities, of referral resources; majorly ambulances or some mode of transportation for sick young infants and caregivers. This results in caregivers having to look for alternative means of transportation to the referral facility and in most cases meant the use of a motor bike.

> *“The mother usually struggles to bring the child to hospital, sometimes on foot if they have to. The mother sometimes asks for help from a bodaboda (motor bike) rider to transport the child to hospital with the promise that she will pay for the transport later when she is able to get some money. Sometimes if the CHV has a motorbike he or she transports both the mother and child to the facility.” [****CHV in Focus Group***]

Health providers and CHVs sometimes looked for strategies to facilitate referral of a sick young infants due to lack of financial resources.

> *“When the mother comes here and is referred to Lodwar, we are asked to look for transport money or pay for fuel for the County ambulance which is also rarely available.” [****CHV in Focus Group***]

The interaction between culture, religion and older men as care determinants was evident in the discussion with CHVs. These are critical contextual issues that should be considered in the development and implementation of community centric interventions.

> *“Some people still have beliefs of their own. For example a parent might tell him or herself that something has been done to their child. The Bible says that if you believe, you will get healed and as such the parent with the sick child may decide to take the child to one of the older men within the village whom they believe has divine power to heal the child by pouring water on them. Sometimes a goat is slaughtered as directed or done by the men believed to have divine powers.” [****CHV in Focus Group***]

##### 1.8. Drivers of Common infections

Poor hygiene was a key especially in breast feeding mothers was associated with infections in young infants. The common infections that were highlighted included malaria, diarrhea, vomiting, and pneumonia.

> *“…. there is a problem of hygiene for breastfeeding mothers. We keep telling mothers to properly wash their breast before they feed the child but they don’t listen. Washing the breast is a problem for most mothers. They just don’t do it. General body hygiene is also a problem.” [****CHV in Focus Group***]

##### 1.9. Challenges experienced in day 4 and 8 follow up

One of the of the most prevalent challenges to follow up is caregiver’s assumption that infants recover following the first course of treatment involving two intramuscular injections of gentamycin. This meant that there was loss to follow up and lack of adherence to the PSBI management protocol.

> *“…. One of the challenges we encounter is when the child is given the first dose of medicine and the parent presumes that the child is cured meaning treatment is not completed.” [****CHV in Focus Group***]

Distance and availability of transport were compounded by the geographical and physical access vulnerabilities in Turkana county. This is evidenced by the narrations by both CHVs and health providers.

> *“They’ll tell us about the distance, about availability of that transport because in Turkana, transportation in terms of matatu is not readily available so they use the motorbike and sometimes they don’t even have that money to hire a motorbike.”* ***[Provider KII***]
>
> *“When the mother is in the mountains, she finds it quite strenuous to walk with a young child down to and from the top of the mountain which is why some mothers are less inclined to bring the child to hospital.” [****CHV in Focus Group***]

Health providers expressed that day 4 and 8 follow up in the facility are deemed difficult due to perceived competing needs and responsibilities that caregivers have at home, describing follow up as “farfetched”. This indicates a need for provider re-training and induction into PSBI management and IMNCI strategies. This can be achieved through on-job trainings, support supervision and continuous medical education (CMEs) sessions.

> *“On my part, the way I see it is that asking the mother to bring back the child on the eighth and fourth days is a farfetched approach because mothers here have tremendous responsibilities at home coupled with the fact that some of them have no sources of income.”*
>
> [***CHV in Focus Group***]

Some providers did not have a clear linkage and working cooperation with CHVs indicated by their preference to schedule follow up and review sessions directly with caregivers.

> *“….sometimes we really don’t have that linkage with the CHVs to make it easy for the follow-up.” [****Provider KII***]
>
> *“We have been doing very well in terms of linkage. There are villages like I’ve said which CHVs are working very well. So that’s why we do normally follow-up with the mother directly.” [****Provider KII***]

##### 1.10. Availability of commodities

Across all six participating facilities, there was a general shortage or an altogether lack of essential commodities required for management of PSBI.

> *“This hospital is currently serving a large number of people and drug shortages are a common occurrence. There is an urgent need to add on the list of drugs available.”* ***[CHV in Focus Group***]

This lack in antibiotics results in alternative care seeking through traditional medicines that sometimes prove harmful to the young infant.

> *“Even though, the ministry of health provides medicines, the hospital runs out of medication and the parents are told to buy at the chemist. If this happens, the parent ignores the medication all the same and resorts to traditional medicine.” [****CHV in Focus Group***]

Some CHVs indicated that they desired an expanded mandate allowing them to manage minor and non-critical ailments.

> *“If we had the kit we would earn the community’s trust because then they would come to us knowing that they will get some form of immediate treatment and if the condition is serious, we would then offer some form of first aid and afterwards immediately refer the patient having identified the danger signs.” [****CHV in Focus Group***]

##### 1.11. Impact of the COVID-19 pandemic on community health seeking

Availability of personal protective equipment and essential medicines was a major challenge experienced during the COVID-19 pandemic affecting the availability of health care services. There was a reduction in the care seeking evidenced by the reduction in people seeking health services in the facilities.

> *“The pandemic has affected care seeking because even the population has reduced. um It’s like people have fear. um And we are not going to see clients without face mask-even the client who does not have face mask maybe in the community, they’re trying to go and look for that face mask.” [****Provider KII***]

The economic ramifications that resulted due to the COVID-19 pandemic negatively impacted the income of caregivers and households in general. This was majorly due to the public health restrictions to contain spread of the disease resulting in closure of businesses and premises where caregivers, especially women worked.

> *“Another is the lack of financial stability. Women here used to earn little money from working in businesses, but ever since the pandemic began and the subsequent shut down of a lot of businesses especially hotels, these women lack income and are forced to manage their kids at home.” [****CHV in Focus Group***]

### 2. Objective 2: Feasibility and adoption of integrated communication strategies

#### 2.1. Experience using PSBI support tools

The health providers and CHVs demonstrated increased confidence in the management of sick young infants as highlighted below.

> *“…*..*when you see a number of clients may be the client has come with a previous condition or illness that you’ve seen from other clients, you will be able to quickly think of treatment that you gave first so that will enable you to-to have that confidence this client has severe pneumonia, he has [Inaudible] difficulty in breathing.” [****Provider KII***]

There was also increased utilization of PSBI support tools with notable reference to the MoH 100 tool for the identification of danger signs by CHVs and the under-five register by facility health providers.

> *“As a CHV I have no medication to give to the child so I write for him or her the MOH 100 and ensure that the child gets to hospital by possibly accompanying the mother all the way to hospital. When the child is back at the village, you still have to follow-up on them and monitor the child’s condition every step of the way.” [****CHV in Focus Group***]
>
> *“I will rush to the child’s location and inspect him or her while filling in my MOH 100. I will then hand the form to the mother and ask her to immediately take the sick child to hospital.” [****CHV in Focus Group***]

The participants made reference to the Integrated community case management (iCCM) guidelines that are geared towards having a unified strengthened community-led approach in the management of community conditions.

> *“As usual, we use the MOH 100 document to execute the referral procedure. There is this one belonging to ICCM. It used to be called Sick Child Recording Form which we used to use but once it came to end, we started using the MOH 100 even though the MOH 100b was originally designed for adults.” [****CHV in Focus Group***]

#### 2.2. Number of sick young infants followed up and reviewed on day 4 and 8 by providers and CHVs

Generally, the trends in follow up and referral for sick young infants with PSBI indicated a significant decline in the numbers in 2020, resulting from COVID-19 pandemic. This is represented in Figure 1 below.

**Fig. 1:**
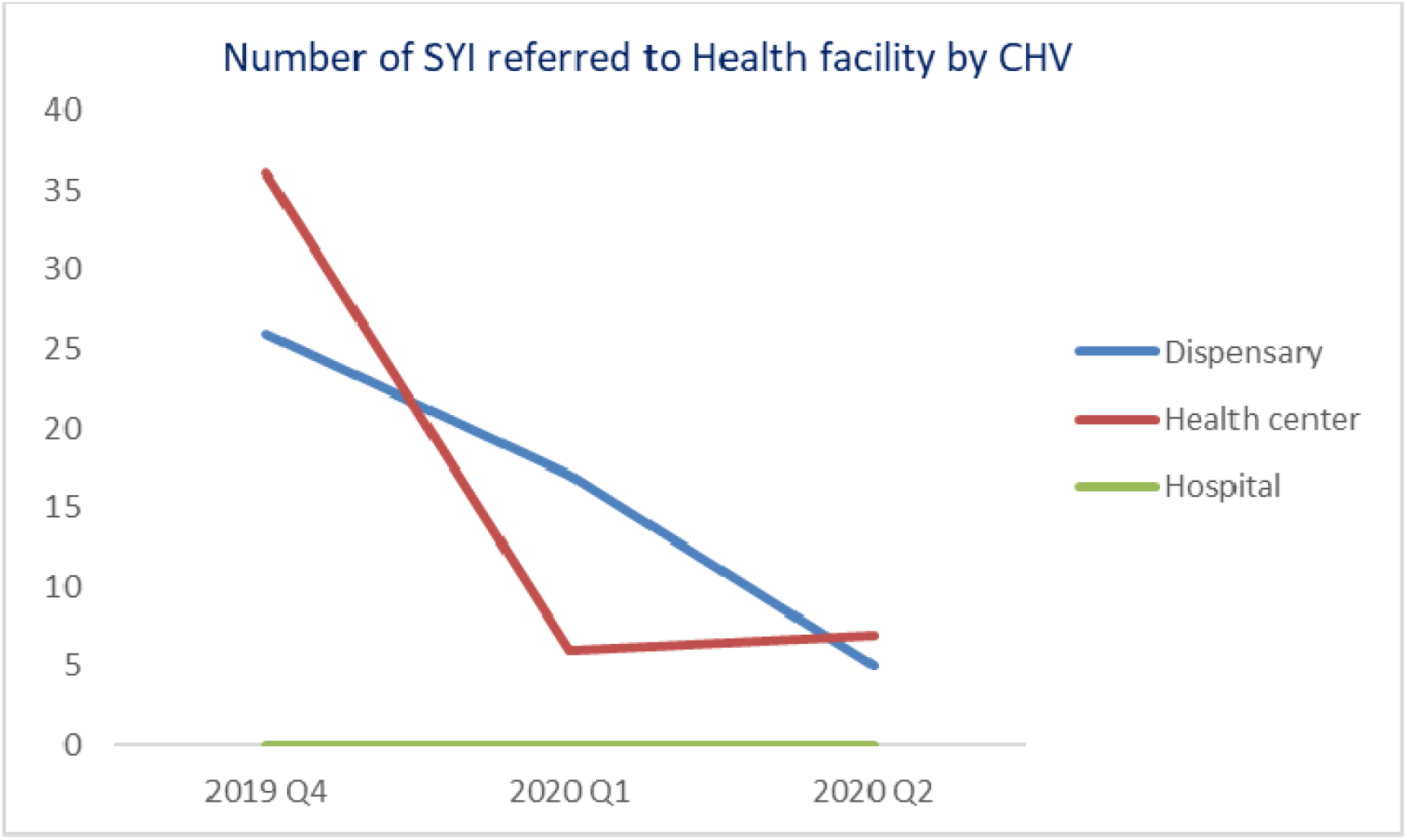
Number of sick young infants referred to the health facility by CHV

The numbers of sick young infants seen on day 4 decreased exponentially in dispensaries compared to a slight decrease witnessed in health centers. This was attributed to closure of most of the dispensaries due to the pandemic. (Figure 2). The numbers in the hospital remained at zero because the sick young infants were managed with inpatient care or did not return on day 4, with qualitative data backing this finding.

**Figure 2:**
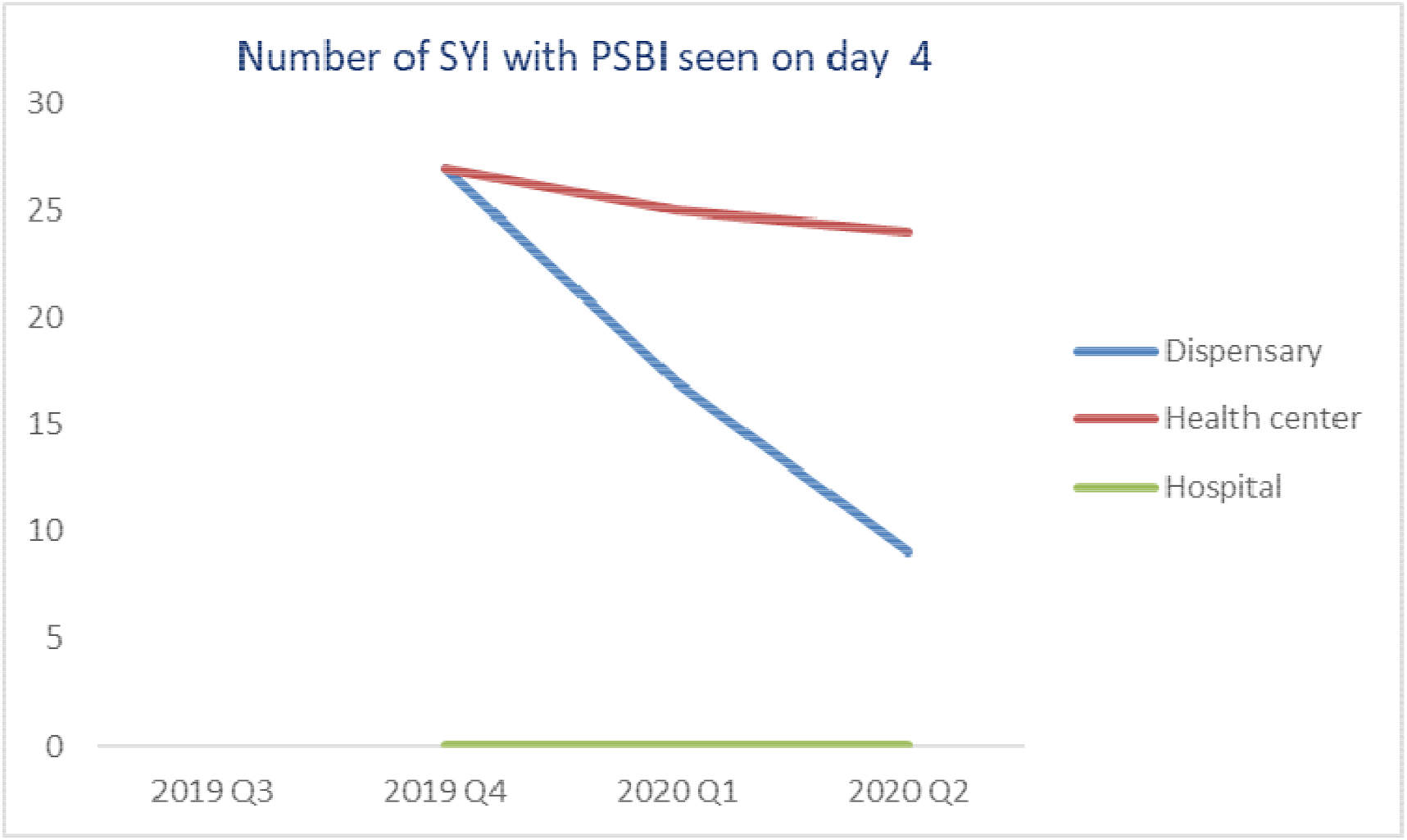
Number of sick young infants with PSBI reviewed on day 4.

> *“The distance they cover to reach here is long. Coming-telling them to come back at day four and the child is stable, they normally don’t come back.” [****Provider KII]***

The number of sick young infants reviewed on day 8 declined in both dispensaries and health centers but increased in health centers at the start of the interventions using strengthened community communication strategies. (Figure 3). This is indicative that improved communication channels between providers and CHVs could potentially improve the number of sick young infants followed up on day 4 and 8.

**Fig. 3:**
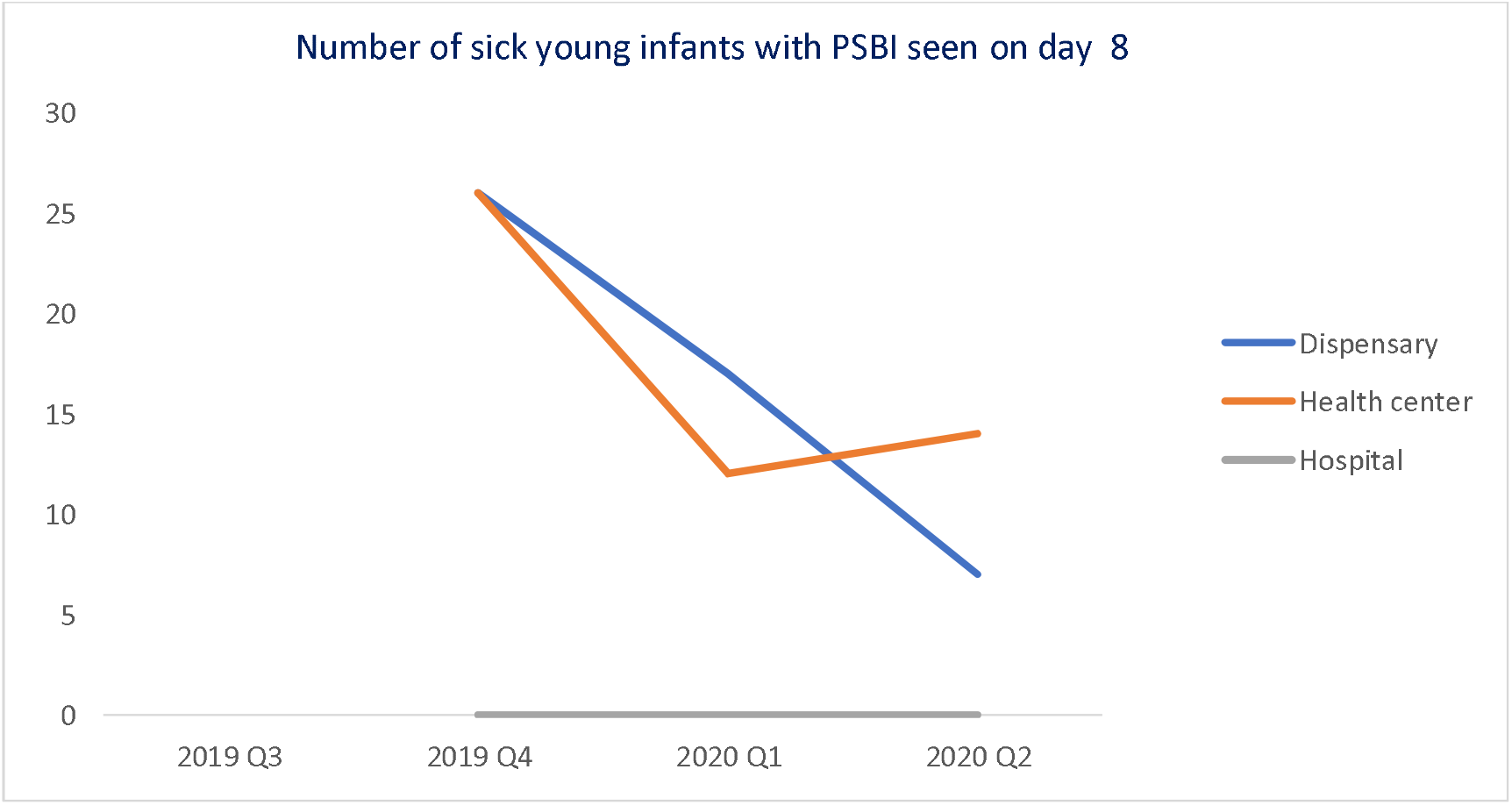
Number of sick young infants with PSBI reviewed on day 8.

#### 2.3. Sick young infants who were reviewed and followed up in line with PSBI guidelines

The number of sick young infants identified with pneumonia or fast breathing in dispensaries was 29 and 8 in the months of August and September 2020 respectively. The number in health centers recorded was 5 for both months. (Figure 4). This was derived from the PSBI assessment and follow up forms available in facilities.

**Fig 4:**
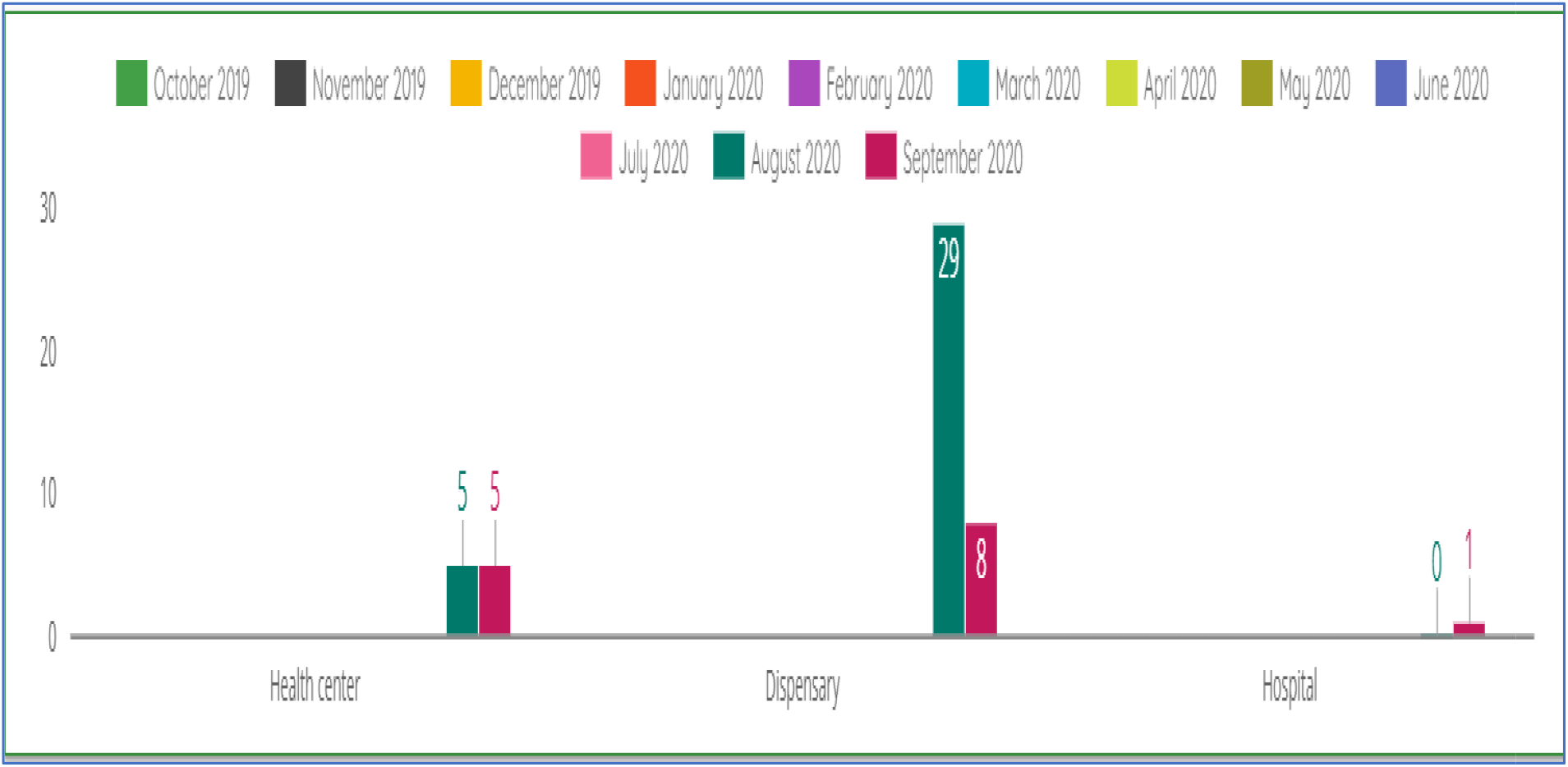
Number of sick young infants identified and treated with Pneumonia/Fast breathing in August and September 2020.

The sick young infants with severe pneumonia ere 11 in dispensaries in August 2020, one in health centers and hospitals for the same month and 3 and 2 for health centers and hospitals respectively in September 2020. There were no sick young infants with pneumonia recorded in dispensaries in September 2020. (Figure 5).

**Fig 5:**
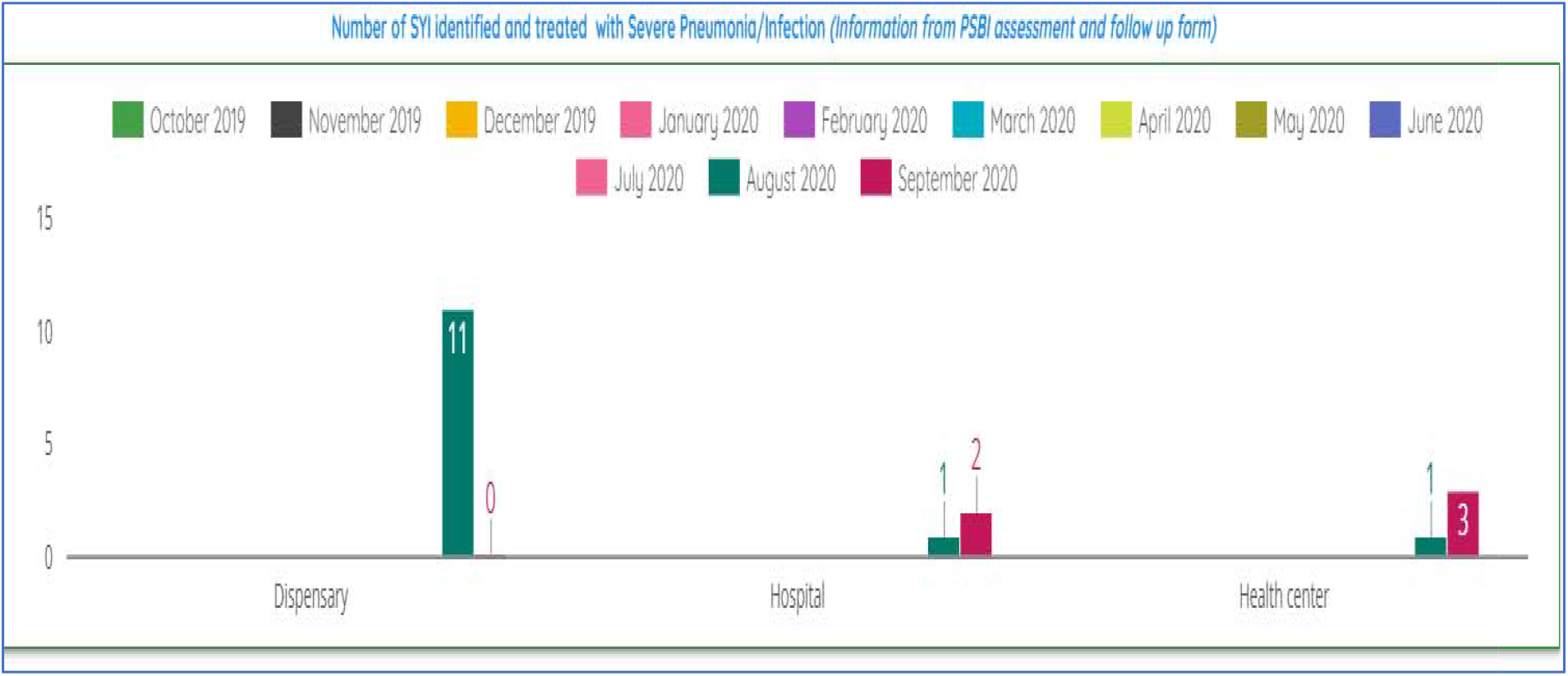
Number of sick young infants identified and treated with Severe Pneumonia/Infection in August and September 2020.

### 2.4. Community referrals of sick young infants by CHVs

16 sick young infants were referred by CHVs to health centers in August and September 2020 while 46 were referred to dispensaries. Regarding PSBI management, 3 sick young infants were documented as referred to health centers by CHVs, and one to dispensaries and hospitals. (Figure 7).

**Fig 6:**
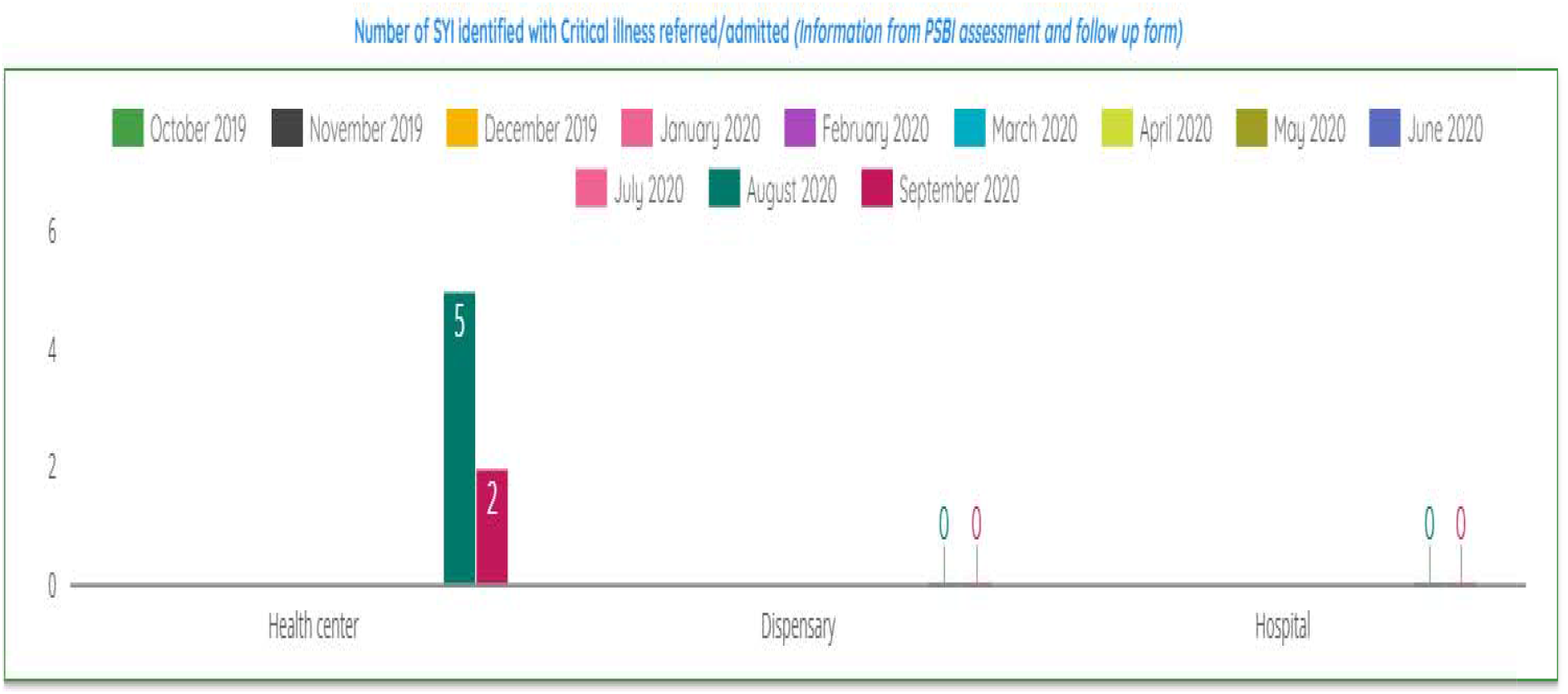
Number of sick young infants with critical illness in August and September 2020.

**Fig 7:**
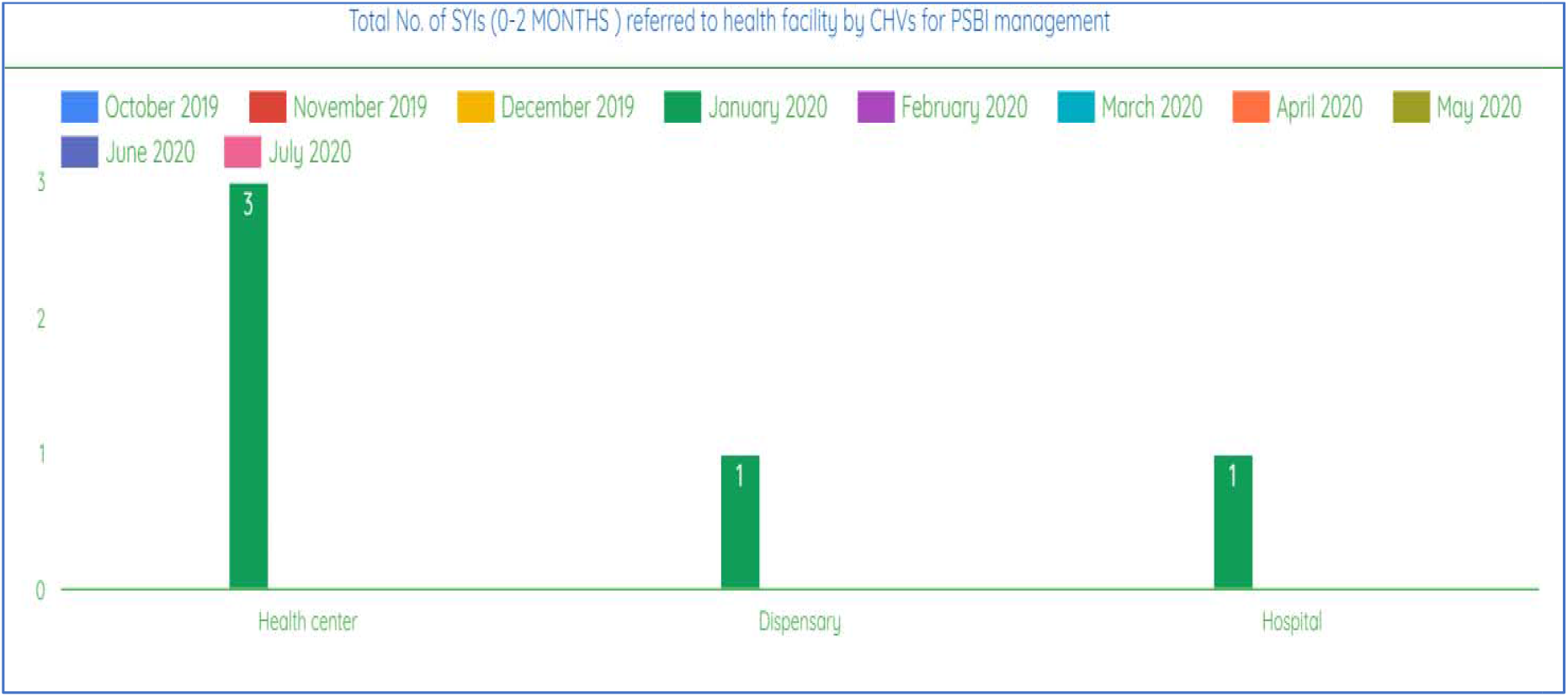
Total number of sick young infants referred to health facilities by CHVs for PSBI management

## Discussion

Community-based approaches have succeeded previously in management of conditions such as tuberculosis and malaria serving as a reasonable testament towards the study of community PSBI management [20]. The available evidence suggests that understanding the barriers and facilitators of community centric approaches in PSBI guidelines implementation is imperative towards promotion of community buy-in and ownership [21]. Competency in identification of PSBI danger signs by CHVs, community engagement forums, regular meetings between CHVs and health providers to strengthen the community system linkage, county support and motivation of CHVs were some of the facilitators of a community-led approach of PSBI management. Positive care seeking to increase demand can be ensured by use of health campaigns involving community members in targeted health messaging [22,23]. Along similar lines, Rupak et al. [24] argues that educational outreach programs should emphasizes the importance home visitation and identification of danger signs by Accredited Social Health Activists, equivalent to CHVs, in the Indian context. In Malawi, Guenther et al. [13] concluded that community sensitization to increase care seeking and joint capacity building of health providers are critical steps in scale up of PSBI implementation.

Poor maternal hygiene lies at the heart of the discussion on drivers of common infections in sick young infants. A consensus view is shared Asfaha et al., [23] where gaps in hygiene practices and unmet maternal nutritional needs were identified as drivers of illness in sick young infants. The barriers to community centric PSBI management include geographical vulnerabilities and accessibility concerns resulting in low referral compliance by caregivers. This is concurrent with findings by Applegate et al. [17] in rural Bangladesh that highlighted caregiver acceptability of PSBI guidelines. Additionally, deficits in referral pathways ipso facto resulted in poor community-facility referral, majorly caused by lack of emergency transport via ambulances. Lack of transportation and distance are key barriers to accessing health care in low resource settings [21,25]. The lack of essential commodities (pharmaceuticals and non-pharmaceuticals) was evident in most facilities leading to negative community trends in care seeking. There is need to ensure timely availability of essential commodities for uninterrupted provision of care and treatment to sick young infants. This is reiterated by Ayede et al. [26] in Nigeria.

A client tracking form and strengthening of communication and facility linkage of CHVs by phone calls and short messaging services (SMS) were introduced to ensure the community management protocol for PSBI is adhered to. Caregivers report fewer delays in health facilities when the community health worker called ahead during referral [17]. Strengthened communication also facilitated community referral of critically illness and severe pneumonia to responsive facilities. The client tracking form served as a routine tracking tool for sick young infants referred by frontline health providers to ensure that they are reviewed by a CHV in the community. Integration of community PSBI indicators in routine monitoring systems is vital to ensure effective monitoring of treatment outcomes [27].

Community engagement and participation is essential in enhancing community-based interventions with their involvement in the conceptualization, design, and implementation of programmatic health care delivery interventions. This augments acceptability [28] of community centric strategies.

## Conclusion

This study seeks to contribute evidence on strengthening PSBI community management by enhancing day 4 and day 8 follow up, review and community referral of sick young infants in Turkana, Kenya. The feasibility, adoption, and fidelity of strengthening community facility linkage through integrated communication strategies was documented, indicative of a successful community-facility linkage in dispensaries and health centers despite the effects of the COVID-19 pandemic. The data will be instrumental in future scale up of integrated community-facility linkage and strengthened follow up of sick young infants on day 4 and day 8.

## Data Availability

All data produced in the present study are available upon reasonable request to the authors.

## Abbreviations

PSBI: Possible Severe Bacterial Infection
WHO: World Health Organization
CHW: Community Health Worker
CHV: Community Health Volunteer
MKU: Mount Kenya University
KEPRECON: Kenya Paediatric Research Consortium
IMNCI: Integrated Management of Newborn and Childhood and Illnesses
CMEs: Continuous Medical Education
iCCM: Integrated Community Case Management

## Acknowledgements

We acknowledge the contribution of the study participants and the Turkana County Health Management Team for their support during this research study. The project that generated data used in this study was made possible by the support of the United Nations Children’s Fund (UNICEF) under the terms of the contract number 43291592.

## Notes

### Competing Interest Statement

The authors have declared no competing interest.

### Author Declarations

Ethical clearance was sought and granted from Mount Kenya University-Institutional Scientific Ethics Review Committee No. MKU/ERC/1596.

