## Supplemental Table 1 for "Lessons learnt from community referral and follow up of sick young infants with Possible Severe Bacterial Infection in Turkana County, Kenya"

**Table 1: Summary of main gaps and suggested solutions for follow up and review of sick young infants with possible severe bacterial infection in Turkana county, Kenya**

| **Main Gap** | **Suggested solution** |
| --- | --- |
| **Barriers to enhanced compliance to follow up on days 4 and 8 and community referral pathways** | |
| Socio-cultural deterrents of skilled birth delivery in a facility | Community dialogue, education and sensitization with community focal persons coupled with creation of champions for skilled delivery will help alleviate misconceptions on facility deliveries. |
| Harmful community socio-cultural practices resulting in PSBI | • Strengthen CHVs as referral agents and improve facility referral systems  • Support CHVs to create demand for prompt care seeking using job aids  • Use CHV to ensure community messaging on common causes of illness tailored towards demystifying causes of illness |
| Barriers to prompt care seeking | • Utilization of local resources e.g. CHVs, TBAs and older women and other localized champions for newborn care to advocate for prompt care for SYIs  • Continued sensitization using Information Education Communication Materials that contain simple message on care of infants targeting barriers to care seeking that can be used by various actors at community level |
| Weak community referral pathway | • County investment in community ambulances to facilitate referral of SYIs  • Appropriate renumeration and motivation for CHVs facilitating referral of SYIs. |
| Challenges in day 4 and day 8 follow up | • Community education on appropriate completion of PSBI treatment  • Strengthened community system by empowerment and investment in CHVs to ensure adherence to day 4 and 8 follow up visits. |
| Stock outs of essential commodities | • Strengthen the forecasting, quantification, procurement, and distribution of essential commodities at the county and sub county levels |
| Decreased care seeking and service delivery interruption due to the COVID pandemic | • Provision of Protective Personal Equipment for health care workers  • Ensuring uninterrupted service provision during the COVID pandemic for continuous service delivery |
